# Prevalence of Soil Transmitted Helminths Among School Children in Aliade, Gwer- East Local Government Area, Benue State, Nigeria

**DOI:** 10.1101/2023.05.15.23290014

**Authors:** Dorcas Asoo Yaji, Agba Jerome Terzungwe, Isegbe Emmanuel Onah

## Abstract

**Objectives:** Soil-transmitted helminthes (STHs) refer to the intestinal worms infecting humans that are transmitted through contaminated soil. STH is endemic in Nigeria. This study determined the prevalence among school aged children in Aliade, Gwer-East Local Government Area of Benue State, Nigeria.

**Setting:** The study was carried out among pupils aged 5-20 years from four schools in the Government Area.

**Participants:** Stool samples were collected from 342 pupils, 196(57.3%) male and 146(42.7%) female and examined for helminth eggs using Direct wet mount method and Formol Ether Concentration technique. Chi square was used to compare the relationship between variables.

**Results:** Overall prevalence of 96(28.1%) was recorded. Prevalence among male 55(28.1%) and female 41(28.1%) indicated no statistical significance(P=0.997). Eggs of three helminthes, *Ascaris lumbricoides*, Hookworm, and *Trichuris trichiura* were observed with prevalence of 18.1%, 9.1% and 1.8% respectively. Prevalence of *A. lumbricoides* among the male pupils (21.9%) was significantly different compared to prevalence among the female (13.0%) (P=0.034). while Hookworm exhibited higher prevalence among female (31.7%) than male (5.6%), this difference was statistically significant. Age group 9-12years had the highest overall prevalence of 32.1% closely followed by group 5-8years (29.4%), while least prevalence of 10.9% was observed in group13-16 years. There was a significant difference in prevalence by age among the pupils (P= 0.036). Playing in soil and eating of unwashed fruits were found to have a relationship with STHs infection among the children. The two factors have a statistically significant difference(P ≤0.05).

**Conclusions:** The overall prevalence of soil-transmitted helminth infections in Aliade, Gwer-East L.G.A is moderate (28.1%). Community health education and good sanitary and hygienic practices are essential in preventing soil-transmitted helminthiasis.

## 1. Introduction

Soil-transmitted helminths (STHs) are a group of neglected tropical diseases that causes the most common infections worldwide and primarily affect marginalized populations in low- and middle-income countries. The three main species that infect people are *Ascaris lumbricoides, Trichuris trichiura* and hookworms^1^ (STH) infections are tagged as one of the most common and neglected infections globally^2^

Parasitic worms are referred to as helminthes because they live and feed on living hosts. They receive both nourishment and protection by disrupting the host’s ability to absorb nutrients, resulting in weakness and disease of the host.^3^ Infected children are nutritionally and physically impaired.^4^ They also cause blockage of the lumen thereby reducing the absorptive surface, feeding on the already digested food of the host, pressure from an enlarging larva, ingestion of blood and disrupting the intestinal mucosa by adult worms which could lead to anaemia and other conditions.^5^ Growth stunting, diminished physical fitness, impaired memory, and cognition are also some health issues associated with STHs infections^6,7^ Infections with hookworms may lead to intestinal blood loss that results in irondeficiency anaemia^8^ Ascariasis can be asymptomatic, causing only malnutrition and growth retardation, or it may present with abdominal pain, nausea, vomiting, bloating, and diarrhea.^9^ Infection with T. trichiura infection may lead to rectal bleeding and abdominal pain.^10^

They are well adapted to the parasitic lifestyle characterized by efficient attachment organs and suckers. Soil-transmitted helminthes have been among the major chronic parasitic infections distributed throughout the world. Globally, about 4.5 billion individuals are at risk and more than 1.5 billion people become infected, of which about 568 million suffer from the infection most been school-age children.^11^

Infections cause by STHs is among the leading causes of global health problems, especially among the poorest and deprived communities where implementation of control measures is difficult.^12^ General occurrence and distribution of STH infections is associated with factors such as socio-economic status of the community, poor sanitation of the environment and lack of awareness about personal hygiene, latrine usage, lack of access to good water, and climatic changes.^11^

Soil-transmitted helminthes live in the intestine and their eggs are passed into faeces of infected persons. If an infected person defecates outside (near bushes, in a garden, or field) or if faeces of an infected person are used as fertilizer, eggs are deposited in soil. Ascaris and hookworm eggs become infective as they mature in soil. People are infected with Ascaris and whipworm (*Trichuris trichiura*) when eggs are ingested. This can happen when hands or fingers that have contaminated dirt on them are put in the mouth or by consuming vegetables and fruits that have not been carefully cooked, washed or peeled. Hookworm eggs are not infective; they hatch in soil releasing larvae (immature worms) that mature into a form that can penetrate the skin of humans. Hookworm infection is transmitted primarily by walking bare-footed on contaminated soil. *Ancylostoma duodenale* can also be transmitted through ingestion of larvae. Light or low infection by soil-transmitted helminthes usually show no symptoms, heavy infections on the other hand causes a range of health problems, including abdominal pain, diarrhea, blood, and protein lose, rectal prolapsed and physical and cognitive growth retardation.^1^ Environmental survival of STH eggs and larvae including hatching and embryonation are determined by warm temperatures and adequate moisture.^12^

For basic diagnosis, specific helminthes can be generally identified from the faeces, and their eggs microscopically examined and enumerated using faecal egg count method. However, there are certain limitations such as the inability to identify mixed infections, and on clinical practice, the technique is inaccurate and unreliable. A novel effective method for egg analysis is the Kato-Katz technique. It is highly accurate and rapid for diagnosis of *A. lumbricoides* and *T. trichiura*; however not so much for hookworm, which could be due to fast degeneration of the rather delicate hookworm eggs.^13^

In Nigeria, a considerable amount of human and animal wastes is discharged into the soil daily, leading to the contamination of the soil with STHs eggs and larvae.^14^ The practice is common among the people of Aliade, Gwer-East Local Government Area of Benue State, Nigeria which predisposes the inhabitants of the area to STHs infection hence the need for this study.

## 2. Materials and Methods

### 2.1 Study area

The study was conducted on school children in Aliade, Gwer-East Local Government Area of Benue State, Nigeria between February to June 2019. Four schools (3) primary and one (1) junior secondary school where randomly selected.

Aliade is located on the coordinates 7°18’0” N 8° 28’60”E. The climate of the area is typically tropical with a characteristic dry season of about six (6) months (October to March) and a wet season of about six (6) months (April to September), the average annual rainfall ranges from 1000-1250m. The area experiences an average humidity is between 75-100% and a mean annual temperature of about 30°C (Wikipedia, 2018). The study area has tropical rainforest vegetation characterized by tall trees and grasses. The inhabitants of Aliade are mainly Tiv speaking people with a mixture of other Nigerian ethnic groups. The people are mostly farmers and the major source of water supply in the area is well and stream.

### 2.2 Sample Size and Sampling Technique

The sample size was calculated using population proportion estimate formula as described by Canhoto^15^ at 95% confidence level and 50% proportion. Using the margin of error of 5% the sample size was calculated as:

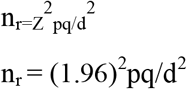

where n_r_ refers to the require sample size, Z a constant (1.96 at 95% confidence level), p represents the proportion having the characteristics (helminth infections), d the degree of precision (acceptable marginal error) in this case 5%, and q=1-p.

### 2.3 Sample Collection

The heads of the schools were first contacted and permission was obtained from them prior to the commencement of the study. Seminars were organized to educate the children on the concept of STH infections. The heads of schools were very instrumental in mobilizing and encouraging their subjects to participate in the study. Children that were willing were given questionnaire to obtain demographic and environmental information and educational knowledge of soil-transmitted helminth infections. Pre-labeled sample containers were then given to each subject to donate stool sample. They were instructed not to contaminate the samples with urine or disinfectants.

### 2.4 Laboratory Method

The specimen containers where retrieved from the children with about 4g of stool sample in each. The specimens where first examined by direct method. Specimens that test negative with direct method were further subjected to formol ether concentration (FEC) technique as described by Cheesbrough,^16^ Both methods are described below.

### 2.4.1. Direct wet mount method

A drop of normal saline (0.90%) and iodine solution was added to each slide separately. With a wire loop a portion of the faecal sample was placed on the slides. The set ups were homogenized and covered with cover slips. Each slide (with two preparations) was then examined under the microscope using X10and X40 objective lens with the condenser iris close enough to give a good contrast. The glass slides were labeled and numbered to avoid treating wrongly. Observations were recorded as type of STH eggs found on the preparation.

### 2.4.2. Concentration technique (Formol Ether Concentration)

For each sample, a gram was placed in the baker and 4ml of 10% formalin was added to it and mixed. Again, 3ml of formalin was added. The solution was sieved to remove large particles and then transferred to into a test tube. 4ml of ethyl acetate was added into the test tube containing the solution and shaken very well. The content was then transferred into centrifuge tubes and inserted into the centrifuge and the machine was run at 3,500revolutions per minute for five minutes. The tube was removed and the supernatant decanted. two smears, one with saline and another with iodine were then prepared from the remaining content, cover slipped and examined microscopically for eggs of soil-transmitted helminthes. Observations were recorded as type of helminthes and number of eggs per gram of stool sample.

### 2.5 Ethical consideration

Ethical clearance to carry out the study was obtained from the Benue State Ministry of Health, Makurdi.

### 2.6 Data Analysis

Data collected through questionnaire and results of laboratory analysis was coded and analyzed using SPSS version 23.0. The prevalence of STH infections calculated and expressed as a percentage of positive cases over the number examined. Chi-square was used to compare the differences in prevalence based on gender, age range, schools sampled and risk factors. Significance was determined at p<0.05.

## 3. Results and Discussion

### 3.1 Demographic characteristics of the study subjects

A total of 342 samples were examined comprising of 196 (57.3%) males and 146 (42.7%) females (table 1). Study subjects fall into one of four groups: 5-8, 9-12, 13-16, 17-20 years. Group 9-12 had the highest number of participants, 159 (46.5) and the least number of participants occurred in group 17-20 having only 1(0.3) participant (table 2).

**Table 1.** Number of participants with regards to sex.

| Sex | Total No examined | Percentage (%) |
| --- | --- | --- |
| Male | 196 | 57.3 |
| Female | 146 | 42.7 |
| **Total** | **342** | **100** |

**Table 2.** Number of participants with regards to age group.

| Age group | Total number examined | Percentage (%) |
| --- | --- | --- |
| 5-8 years | 136 | 39.8 |
| 9-12 years | 159 | 46.5 |
| 13-16 years | 46 | 13.5 |
| 17-20 years | 1 | 0.3 |
| **Total** | **342** | **100** |

### 3.2 Prevalence of STHs infections based on age group

study subjects where group into four groups with an interval of four starting from pupils of age five and above. Highest prevalence of 54(32.1%) occurred in children within the age range 9-12 followed by children within the age range 5-8; 43(31.6%) and then age group 13-16; 5(10.9%). Age group 17-20 had only one participant with a prevalence of 0.0% (Table 3). The overall prevalence showed a statistical significance in relation to age groups(P=0.036). independent infection with parasites showed no statistical significance in the prevalence based on the age groups (p>0.05). *A. lumbricoides* had the highest prevalence among pupils of age group 5-8; 27(19.9%), closely followed by age group 9-12; 31(19.5%) while age group 13-16 had the least prevalence of 4(8.7%). Age group 5-8 also had the highest cases of Hookworm infection, 14(10.3%), this was closely followed by age group 9-12 which had a prevalence 16(10.1%). Age group 13-16 had only a single case of Hookworm infection representing 2.2% of the persons sampled under this group (table 3). *Trichuris trichiura* infection occurred only in members of age group 9-12 and age group 5-8 with prevalence of 4(2.5%) and 2(1.5%) respectively.

**Table 3.** prevalence of soil-transmitted helminths based on age groups.

| Age group | Total number examined | Prevalence |  |  |  |
| --- | --- | --- | --- | --- | --- |
|  |  | Overall prevalence | *Ascaris lumbricoides* (%) | Hookworm (%) | *Trichuris trichiura* (%) |
| 5-8 | 136 | 40(29.4) | 27(19.9) | 14(10.3) | 2(1.5) |
| 9-12 | 159 | 51(32.1) | 31(19.5) | 16(10.1) | 4(2.5) |
| 13-16 | 46 | 5(10.9) | 4(8.7) | 1(2.2) | 0(0.0) |
| 17-20 | 1 | 0(0.0) | 0(0.0) | 0(0.0) | 0(0.0) |
| Total | 342 | 96(28.1) | 62(18.1) | 31(9.1) | 6(1.8) |
| Chi square |  | 8.515<sup>a</sup> | 3.452<sup>a</sup> | 3.191<sup>a</sup> | 1.438<sup>a</sup> |
| Df |  | 3 | 3 | 3 | 3 |
| P value |  | 0.036* | 0.327ns | 0.363ns | 0.697ns |

Significant (\*) at $P\leq 0.05$ , Not Significant (ns) at $p>0.05$

### 3.3 Prevalence of STHs infection based on Sex

A total of 342 samples were examined, 196(57.3%) male and 146(42.7%) female. The overall prevalence of STHs infections among sexes was 55(28.1%) male and 41(28.1%) female having a total STHs infection prevalence of 96(28.1%) prevalence in relation to age disparity was not statistically significant(P>0.05) (table 4).

**Table 4.** prevalence of STHs infections based on sex.

| Sex | Total number examined | Prevalence |  |  |  |
| --- | --- | --- | --- | --- | --- |
|  |  | Overall prevalence (%) | *Ascaris lumbricoides* (%) | Hookworm (%) | *Trichuris trichiura* (%) |
| Male | 196 | 55(28.1) | 43(21.9) | 11(5.6) | 4(2.0) |
| Female | 146 | 41(28.1) | 19(13.0) | 20(13.7) | 2(1.4) |
| Total | 342 | 96(28.1) | 62(18.1) | 31(9.1) | 6(1.8) |
| Chi square |  | 0.000 | 4.491 | 6.638 | 0.219 |
| Df |  | 1 | 1 | 1 | 1 |
| P value |  | 0.997ns | 0.034* | 0.010* | 0.640ns |

Significant (\*) at $P \leq 0.05$ , Not Significant (ns) at $p > 0.05$

Infections with *A. lumbricoides* was more prevalent in males 43(21.9%) than females 19(13.0%) while Hookworm infections exhibited more prevalence in females 20(13.7%) than in males 11(5.6%). The differences in the prevalence of *A. lumbricoides* and hookworm with respect to sexes of the study subjects were statistically significant (P≤0.05). *T. trichiura* infection was more prevalent in male 4(2.0%) than in female 2(1.4%), but the difference was not statistically significant (table 4).

### 3.4 Prevalence of STHs based on school

The number of samples collected from the schools were, School A, 80(23.4%), School B 89(26.0%), School C 41 (12.0%) and School C 132 (38.6%) (table 5).

**Table 5.**
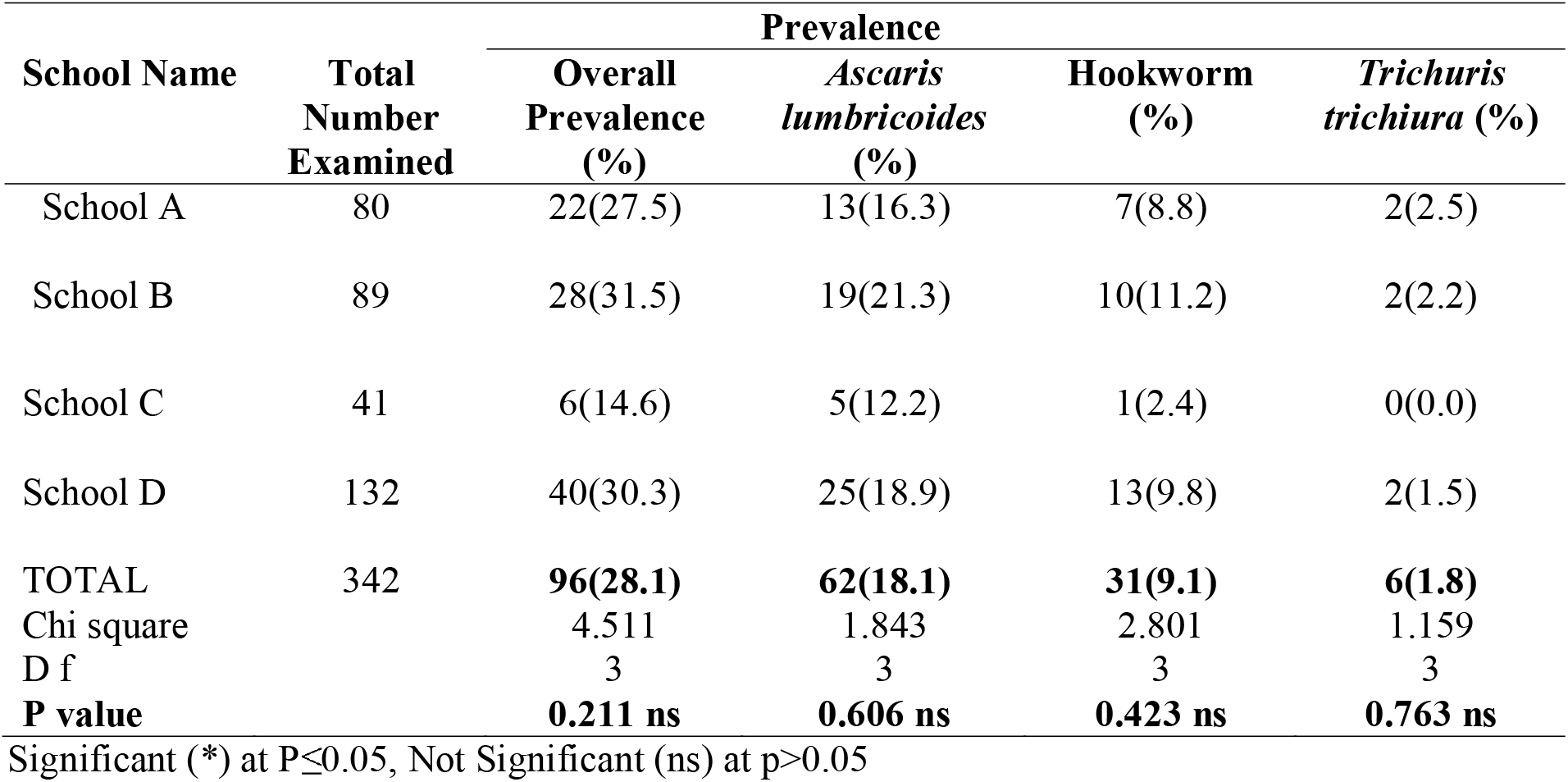
Prevalence of STHs infections based on school

| School Name | Total Number Examined | Prevalence |  |  |  |
| --- | --- | --- | --- | --- | --- |
|  |  | Overall Prevalence (%) | *Ascaris lumbricoides* (%) | Hookworm (%) | *Trichuris trichiura* (%) |
| School A | 80 | 22(27.5) | 13(16.3) | 7(8.8) | 2(2.5) |
| School B | 89 | 28(31.5) | 19(21.3) | 10(11.2) | 2(2.2) |
| School C | 41 | 6(14.6) | 5(12.2) | 1(2.4) | 0(0.0) |
| School D | 132 | 40(30.3) | 25(18.9) | 13(9.8) | 2(1.5) |
| TOTAL | 342 | **96(28.1)** | **62(18.1)** | **31(9.1)** | **6(1.8)** |
| Chi square |  | 4.511 | 1.843 | 2.801 | 1.159 |
| D f |  | 3 | 3 | 3 | 3 |
| **P value** |  | **0.211 ns** | **0.606 ns** | **0.423 ns** | **0.763 ns** |

Significant (\*) at $P \leq 0.05$ , Not Significant (ns) at $p > 0.05$

School B had the highest prevalence of soil-transmitted helminthic infection of 28 pupils representing 31.5% of the subjects in the school followed by School D, 40(30.3%), and school A, 22(27.5%), while school C had the least prevalence with 6 subjects infected representing 14.6% of the participants in the school, (table 5).

School B had the highest STHs infection prevalence of 19(21.3%) and 10(11.2%) for *Ascaris lumbricoides* and Hookworms respectively followed by school D, having STHs infection prevalence of 25(18.9%) and 13(9.8%) for *A. lumbricoides* and Hookworm respectively. School A ranked third with prevalence of 13(16.3%) *A. lumbricoides* 7(8.8%) Hookworm and had the highest prevalence of *Trichuris trichiura*2(2.5%). While school C had the least prevalence of *A. lumbricoides* 5(12.2%), Hookworm 1(2.4%) and 0.0% prevalence for *T. trichiura*.

### 3.5 Prevalence of soil-transmitted helminths infection based on risk factors

with regards to *A. lumbricoides* infection, analysis of data collected through questionnaire reviewed that people who sometimes play in soil have higher prevalence 49(24.7%) than those who do not 13(9.0%) with a significant difference, that is P≤0.05 (table 6). Similarly, higher prevalence 43(27.6%) was observed from people who do not always wash their fruits before eating as compare to people who do, 19(10.2%). This relatedness was statistically significant (P≤0.05)

**Table 6.** prevalence of soil-transmitted helminths infections based on risk factors as reported via questionnaire.

| Risk factor |  | Status |  | Chi square | P value. |
| --- | --- | --- | --- | --- | --- |
|  |  | Infected (+) | Not infected (-) |  |  |
| Do you sometimes play in the soil? | Yes | 49(24.7) | 149(75.3) | 13.880<sup>a</sup> | 0.000* |
|  | No | 13(9.0) | 131(91.0) |  |  |
| Do people defecate in the bush around your community? | Yes | 42(19.6) | 172(80.4) | 0.864<sup>a</sup> | 0.353ns |
|  | No | 20(15.6) | 108(84.4) |  |  |
| Is there a toilet facility in your school? | Yes | 38(18.4) | 169(81.6) | 0.005<sup>a</sup> | 0.942ns |
|  | No | 24(18.0) | 109(82.0) |  |  |
| Is there a toilet facility in your house? | Yes | 51(18.0) | 233(82.0) | 0.119a | 0.730ns |
|  | No | 10(20.0) | 40(80) |  |  |
| Do you always wash your fruits before eating? | Yes | 19(10.2) | 167(89.8) | 17.205<sup>a</sup> | 0.000* |
|  | No | 43(27.6) | 113(72.4) |  |  |

Significant (\*) at $P \leq 0.05$ , Not Significant (ns) at $p > 0.05$

Subjects who agreed that cases of defecating in the bush exist in their communities had the highest infection with *A. lumbricoides* 42(19.6%) as compared to subjects who did not agree, 20(15.6%), but the difference was not significant (p>0.05).

Pupils who had toilet facilities in their schools had higher infections of STHs 38(18.4%) than those who had no toilet facilities in their schools. This relatedness was however not statistically significant (p>0.05). The relationship between having a toilet at home or not was not significant (p>0.05), as those who had toilet in their houses had higher prevalence 51(18.0%) than those who had no toilets in their houses 10(20%) (table 6).

### 3.6 Knowledge of the study subjects on the existence of soil-transmitted helminthic infection

Data collected through questionnaire revealed that 176(51.5%) of the participants were aware of the existence of intestinal worms while another large number of participants 166(48.5%) where not aware of the existence of intestinal worms.(Figure one)

## 4. Discussion

The result of this study revealed that the prevalence of soil-transmitted helminths among school children in Aliade was 28.1%. This classifies the community as a moderate risk area for preventive therapy by WHO standard.^15^ Soil-transmitted helminthic infection is therefore a problem among school children in Aliade, Benue State. This report is one of the first findings on STH infection in the study area. The reported prevalence of STH in this study is slightly lower than that reported in Tanzania (29.0%).^17^ and 32.3%, 33.2% and 36.0% in Ethopia, 78.8% in Myanmar.^18,19,20,21^

In contrast, the current STH infections prevalence was higher than the studies conducted in different countries of the world, such as reports from African countries Kenya (5.6%), ^22^ Gabon (15.0%)^23^, Togo (5.0%)^24^, Colombia (29.6%)^25^, India (7.7% and 7.0%)^26,27^ and Thailand (3.13%)^28^ and Nigeria (11.27%)^29^. The possible reason for this inconsistency in the prevalence of STH infections might be due to the differences in socio-cultural determinants, behavioral characteristics, climatic conditions, implementation of prevention and control measures, and frequency and application of mass drug administration on intestinal parasites among different countries.

Prevalence among male and female showed no significant difference. This may be attributed to the general hygienic and sanitary condition of the environment owing to food, water and soil contamination. The non-significance in the study with regards to sex disparity is in line with the report by Emana *et al*.,^30^ but does not agree with the reports by Odinaka *et al*.,^31^ where higher prevalence has been reported to occur in male than female and Olaniran *et al*.,^3^ where higher prevalesnce was recorded in female than male in studies carried out at Central Local Government Area, Ile-Ife, Osun State.

The most prevalent parasite found was *Ascaris lumbricoides* (table 4). This could be due to favourable environmental conditions and poor hand washing habit among the studied subjects. High fecundity of *A. lumbricoides* could be a reason the eggs where encountered in large numbers during the study. The higher prevalence of *A. lumbricoides* relative to Hookworm and *T. trichiura* as shown by this study is in consonance with the work of Ojorungbe *et al*.,^32^.

Hookworm infection was higher in female than male, this is in contrast to the reports were he reported higher prevalence of Hookworm in male in studies carried out on 395 pupils in Ile-Ife, Osun State.^3^ Higher prevalence of Hookworm in female as demonstrated by this study may be as a result of geographical variations.

The randomly selected schools and number of samples collected are school A (80), school B (89), school C (41), and school D (132). Soil-transmitted helminthes infections occurred among some children in all the schools. The result therefore confirms that soil-transmitted helminthic infection is still a problem among Nigerian school children.^33,34^

Age related pattern of infection in the study indicated that age group 9-12 years had the highest overall prevalence for the three STH parasites recovered. However, infections of *Ascaris* and Hookworm were more prevalent among children of age group 5-8 years followed by age group 9-12 years while group 13-16 years had the least infection prevalence of all three STH parasites recovered. Higher prevalence among group 5-8 years and group 9-12 years could be as aresult of poor hand washing habit, playing in soil and eating of unwashed fruits which is usually more common among children of these ages. Decrease in the prevalence from age group 13-16 years may be as a result of more careful sanitary and hygienic habits among participants of this group owing to better knowledge. Higher prevalence of infections among children of age 9-12 years as shown by this study is in line with the work of Salawu and Ughele,^34^.

Questionnaire revealed that 48.5% of the children were not aware of even the existence of STHs, this is most likely due to poor or lack of community health education in the area.

With regards to risk factors, playing in soil and not washing fruits before eating have been incriminated by this study to be related with the transmission of these parasites. Relationship with these factors and infections of *A. lumbricoides* had a significant difference(P=0.000). this agrees with the worked Ojja *et al*.,^35^ Infections were more common among children who had toilet facilities in their school than those who did not. A similar trend was observed in having a toilet at home, as infection was higher among children who said they have toilet facilities at home. Infection of STHs is therefore not based on having a toilet facility, other factors may have been responsible for the prevalence recorded among the children.

## 4.1 Conclusion

The overall prevalence of STH infections in Aliade, Gwer-East L.G.A is moderate (28.1%). The individual species prevalence for the three STHs observed in the study is; *A. lumbricoides* 62(18.1%), Hookworm 31(9.1%) and *T. trichiura* 6(1.8%). STH infections in the study was found to occur equally among male (28.1%) and females (28.1%). Age group 9-12 years had the highest overall prevalence (32.1%). Risk factors such as washing of fruits before eating and playing in soil have significant impact on the prevalence of STHs. Many children are not aware of the existence of STH and as such this predisposes them to STHs infections.

## 4.2.1 Recommendations

The implication of the result of this study is that STH infection is still a problem in Nigeria. To ensure good physical and mental development among children and even adults, The following are recommended: A well-organized health education programme on personal hygiene and community health. Mass treatment is desirable and recommended in Gwer-East Local Government Area, Benue State. Good personal hygiene like washing of fruits and vegetables properly before eating. Basic Water, Sanitation and Hygiene (WASH) infrastructures should be provided in elementary schools to minimize open defecation which facilitate the prevalence of STHs.

## Data Availability

All data produced in the present study are available upon reasonable request to the authors

## Acknowledgments

Thank you very much to the Benue state ministry of Education for granting the researches permission to conduct this study. All the pupils who participated in the study are duly acknowledged.

## Ethics approval

The study protocol was approved by the ethics committee of the Department of Biological Science, Benue state University. Each study participant gave informed consent.

## Data sharing statement

No additional data are available.

## Patient and Public Involvement

Not required.

